# Chasing SARS-CoV-2 XBB.1.16 Recombinant Lineage in India and the Clinical Profile of XBB.1.16 cases in Maharashtra, India

**DOI:** 10.1101/2023.04.22.23288965

**Authors:** Rajesh P. Karyakarte, Rashmita Das, Mansi V. Rajmane, Sonali Dudhate, Jeanne Agarasen, Praveena Pillai, Priyanka M. Chandankhede, Rutika S. Labhshetwar, Yogita Gadiyal, Preeti P. Kulkarni, Safanah Nizarudeen, Suvarna Joshi, Krishanpal Karmodiya, Varsha Potdar

## Abstract

**Background:** SARS-CoV-2 has evolved rapidly, resulting in emergence of lineages with competitive advantage over one another. Co-infections with different SARS-CoV-2 lineages can give rise to recombinant lineages. To date, XBB lineage is the most widespread recombinant lineage worldwide, with the recently named XBB.1.16 lineage causing a surge in the number of COVID-19 cases in India.

**Methodology:** The present study involved retrieval of SARS-CoV-2 genome sequences from India (between 1^st^ December 2022 and 8^th^ April 2023) through GISAID; sequences were curated, followed by lineage and phylogenetic analysis. Demographic and clinical data from Maharashtra, India were collected telephonically, recorded in Microsoft® Excel, and analysed using IBM® SPSS statistics, version 29.0.0.0 (241).

**Results:** A total of 2,944 sequences were downloaded from the GISAID database, of which 2,856 were included in the study following data curation. The sequences from India were dominated by the XBB.1.16* lineage (36.17%) followed by XBB.2.3* (12.11%) and XBB.1.5* (10.36%). Of the 2,856 cases, 693 were from Maharashtra; 386 of these were included in the clinical study. The clinical features of COVID-19 cases with XBB.1.16* infection (XBB.1.16* cases, 276 in number) showed that 92% of those had a symptomatic disease, with fever (67%), cough (42%), rhinorrhoea (33.7%), body ache (14.5%) and fatigue (14.1%) being the most common symptoms. Presence of comorbidity was found in 17.7% of the XBB.1.16* cases. Among the XBB.1.16* cases, 91.7% were vaccinated with at least one dose of vaccine against COVID-19. While 74.3% of XBB.1.16* cases were home-isolated; 25.7% needed hospitalization/institutional quarantine, of these, 33.8% needed oxygen therapy. Out of 276 XBB.1.16* cases, seven (2.5%) cases succumbed to the disease. Majority of XBB.1.16* cases who died belonged to an elderly age group (60 years and above), had underlying comorbid condition/s, and needed supplemental oxygen therapy. The clinical features of COVID-19 cases infected with other co-circulating Omicron variants were similar to XBB.1.16* cases.

**Conclusion:** The study reveals that XBB.1.16* lineage has become the most predominant SARS-CoV-2 lineage in India. The study also shows that the clinical features and outcome of XBB.1.16* cases were similar to those of other co-circulating Omicron lineage infected cases in Maharashtra, India.

## 1. INTRODUCTION

The SARS-CoV-2 has undergone rapid evolution since November 2019, resulting in the emergence of competing lineages, of these 2779 have been designated by pango-designation project (https://github.com/cov-lineages/pango-designation) [1]. SARS-CoV-2 lineages with D614G mutation, replaced the index SARS-CoV-2 lineage globally by July 2020. Later, the B.1.351 lineage (Beta variant) became dominant to be soon replaced by the B.1.617.2 lineage (Delta variant), which was in turn replaced by a more transmissible and immune evasive lineage, the B.1.1.529 lineage (Omicron variant) [2]. This global trend was also seen in India, where three major COVID-19 waves were caused by the index SARS-CoV-2 lineage, the Delta variant, and the Omicron variant [3]. Thus, lineages with a competitive advantage dominated and influenced the transmission dynamics of the SARS-CoV-2 pandemic [2]. Now, during the current period of Omicron variant domination, with varying range of host immunity following earlier infections or vaccinations, an Omicron lineage soup has emerged with 923 non-recombinant Omicron lineages, with periodic surges by different Omicron lineages predominating in distinct geographical locations {the entire list #115 Designated SARS-CoV-2 Non-Recombinant (Sub-)Lineages, Lineage only; Maintained by Andrew Urquhart is available at https://cov-spectrum.org/collections/115 [covSPECTRUM(cov-spectrum.org)]} [4].

Further, co-infection with multiple SARS-CoV-2 lineages has led to evolution of SARS-CoV-2 recombinant lineages. There are 251 recombinant lineages and sub-lineages designated by pango-designation project {the entire list #117 Designated SARS-CoV-2 Recombinant (Sub-)Lineages, Lineage only; Maintained by Andrew Urquhart is available at https://cov-spectrum.org/collections/117 [covSPECTRUM(cov-spectrum.org)]}, of which XBB is the most widespread recombinant lineage to date [5]. The XBB lineage emerged following recombination of two co-circulating BA.2 Omicron sub-lineages, BJ.1 and BM.1.1.1, with a breakpoint between 22,897 and 22,941 positions in the RBD of the spike protein (corresponding to amino acid positions 445-460). It is suggested that XBB evolved somewhere around the Indian subcontinent, later spreading to the world [6]. During February 2023, the major XBB sub-lineages circulating in India were XBB.1, XBB.2, and XBB.1.5 [7].

In mid-February 2023, a rise in number of COVID-19 cases was seen in India [8]. In the samples from the state of Maharashtra, India, XBB.1 and XBB.1.5, were the dominant sub-lineages found in our sequencing laboratory. At the same time, there was an increase in SARS-CoV-2 load in sewage samples from Pune, a city in Maharashtra, India, which indicated a rise in COVID-19 cases [Routine official communication, Council of Scientific and Industrial Research-National Chemical Laboratory (CSIR-NCL), Pune, waste-water surveillance, supported by CSIR, New Delhi, Pune Knowledge Cluster Foundation (PKCF), Pune, and The Rockefeller Foundation, US].

It was perplexing that the pre-existing lineages in circulation were causing a new surge in COVID-19 cases. However, on 4^th^ March 2023, a group of international scientists monitoring the evolution of the SARS-CoV-2 virus discussed the global spread of a saltation lineage, XBB.1 sub-lineage, with the spike (S) mutations E180V, K478R, and S486P on GitHub (https://github.com/cov-lineages/pango-designation/issues/1723). They noticed that this sub-lineage had a significant growth advantage over other lineages. The non-Indian sequences submitted on GISAID indicated a travel history mainly from India. The scientists called for rapid monitoring of the lineage’s growth rate and suggested an early lineage designation. On 5^th^ March 2023, it was designated as Pango lineage XBB.1.16 [9]. Since the lineage was newly designated, it was identified as XBB.1/XBB.1.5 and clade 22F by the bioinformatic pipelines currently used for lineage analysis, as they were not updated then. Therefore, manual analysis for the presence of XBB.1.16 defining mutations was carried out in the genomic sequences of SARS-CoV-2 from Maharashtra that were sequenced in our sequencing laboratory. This manual analysis showed the presence of the defining mutations of XBB.1.16 in those sequences. Presence of this new, fitter, and immune-evasive XBB.1.16 lineage clarified the reason of the sudden surge in number of COVID-19 cases in Maharashtra as well as India.

This study describes the current situation of the XBB.1.16 lineage in India. The study also does a comparative evaluation of the clinical characteristics and outcomes of XBB.1.16 cases and other co-circulating Omicron lineage infected cases in Maharashtra.

## 2. MATERIAL AND METHODS

The present study was conducted as a part of the Indian SARS-CoV-2 Genomics Consortium (INSACOG) sequencing activity in Maharashtra to study the evolution of the SARS-CoV-2 virus and its epidemiological trends. The study protocol for SARS-CoV-2 whole genome sequencing was reviewed and approved by the Institutional Ethics Committee at Byramjee Jeejeebhoy Government Medical College (BJGMC), Pune, Maharashtra, India.

### 2.1. SARS-CoV-2 Whole Genome Sequences from India

To study the first appearance and the epidemiological trends of the XBB.1.16 lineage in India, complete genome sequences of SARS-CoV-2 virus, from 1^st^ December 2022 to 8^th^ April 2023, deposited from different States and Union Territories of India, were retrieved from the GISAID database [10]. The associated metadata was downloaded and used for curation. Entries with complete geographic locations and sample collection dates were included in the study. The findings of this study are based on metadata associated with 2,944 sequences available on GISAID between 1^st^ December 2022 to 8^th^ April 2023, and accessible at epicov.org/epi3/epi_set/230419bg?main=true (Supplemental table 1).

### 2.2. SARS-CoV-2 Lineage and Phylogenetic Analysis

Lineage analysis of the retrieved sequences was done using Phylogenetic Assignment of Named Global Outbreak LINeages (PangoLIN) COVID-19 lineage assigner, version v4.2, pangolin-data version v1.19 (https://pangolin.cog-uk.io/) [11], Nextclade vercel software, version 2.6.1 (preview version) [[Nextclade(nextclade-git-feat-composite-fitness-nextstrain.vercel.app)](https://nextclade-git-feat-composite-fitness-nextstrain.vercel.app/results)] [12] and Ultrafast Sample placement on Existing tRee (UShER), University of California, Santa Cruz (https://genome.ucsc.edu/cgi-bin/hgPhyloPlace) [13]. The maximum likelihood phylogenetic tree was estimated, and tree was constructed using Nextclade Augur and rooted to Wuhan-Hu-1/2019 (MN908947) (https://clades.nextstrain.org/) [12]. Trees were visualized and explored using Auspice version 2.45.2 (https://auspice.us/) [14].

### 2.3. Demographic and Clinical Data Collection of SARS-CoV-2 positive Cases in Maharashtra

Demographic data, including the patient’s age, sex, area of residence, contact number and date of specimen collection and testing, were collected from the metadata files submitted by the RT-PCR laboratories to the sequencing laboratories and the ICMR-COVID-19 Data portal using the unique identification number (ICMR ID). Telephonic interviews with each patient were useful in obtaining clinical details regarding the presence of any symptoms at the time of acute infection, type of isolation required, hospitalization, oxygen requirement, treatment given, and vaccination status. Patients not willing to share their clinical history during the interview were documented and excluded from the study.

### 2.4. Statistical analysis

All demographic and clinical data were recorded using Microsoft® Excel, and analysis was performed using Microsoft® Excel and IBM® SPSS statistics, version 29.0.0.0 (241). The continuous variables were presented as the median and interquartile range (IQR). The Kruskal-Wallis test was used to compare the median values between the lineages. The categorical variables were presented as numbers and percentages. The chi-square test was used to compare categorical variables between the lineages. Fisher’s exact test compared the categorical values with limited data. A *p*-value ≤ 0.05 was considered statistically significant.

## 3. RESULTS

### 3.1. Distribution of SARS-CoV-2 Lineages in India

From 1^st^ December 2022 to 8^th^ April 2023, a total of 2,944 sequences were retrieved from the GISAID database, and 2,856 sequences were included in the study following data curation. A total of 225 different lineages were identified in our dataset following Nextclade Pangolin nomenclature, of which XBB* was the most common lineage identified (79.87%) followed by BQ.1* (6.37%) and BA.2.75* (5.50%) (*indicates sub-lineage/s of that lineage) (***Table 1***).

**Table 1:** Distribution of SARS-CoV-2 lineages among sequences deposited on GISAID from India

| SARS-CoV-2 Lineage Distribution |  | Total Count (Percentage) |  |
| --- | --- | --- | --- |
| XBB* | XBB.1.16 | 2281<br>(79.87%) | 723 (31.70%) |
|  | XBB.1.16.1 |  | 310 (13.59%) |
|  | XBB.2.3 |  | 256 (11.22%) |
|  | XBB.1.5 |  | 197 (8.64%) |
|  | XBB.1 |  | 151 (6.62%) |
|  | XBB.2 |  | 126 (5.52%) |
|  | XBB |  | 80 (3.51%) |
|  | XBB.2.3.2 |  | 54 (2.37%) |
|  | XBB.1.9.1 |  | 51 (2.24%) |
|  | XBB.1.5.28 |  | 44 (1.93%) |
|  | XBB.3 |  | 41 (1.80%) |
|  | XBB.1.9.2 |  | 30 (1.32%) |
|  | XBB.2.5 |  | 25 (1.10%) |
|  | XBB.2.4 |  | 21 (0.92%) |
|  | XBB.2.3.3 |  | 17 (0.75%) |
|  | XBB.2.3.4 |  | 17 (0.75%) |
|  | FL.2 |  | 13 (0.57%) |
|  | XBB.1.5.24 |  | 10 (0.44%) |
|  | XBB.1.9 |  | 9 (0.39%) |
|  | XBB.1.5.18 |  | 8 (0.35%) |
|  | XBB.1.5.12 |  | 6 (0.26%) |
|  | XBB.2.6 |  | 6 (0.26%) |
|  | XBB.1.5.15 |  | 5 (0.22%) |
|  | XBB.1.5.7 |  | 5 (0.22%) |
|  | XBB.8 |  | 5 (0.22%) |
|  | XBB.1.11.1 |  | 4 (0.18%) |
|  | XBB.1.9.3 |  | 4 (0.18%) |
|  | XBB.2.7 |  | 4 (0.18%) |
|  | XBB.1.15 |  | 3 (0.13%) |
|  | XBB.1.22 |  | 3 (0.13%) |
|  | XBB.1.3 |  | 3 (0.13%) |
|  | XBB.1.5.13 |  | 3 (0.13%) |
|  | XBB.1.5.17 |  | 3 (0.13%) |
|  | EG.1 |  | 3 (0.13%) |
|  | FL.1 |  | 3 (0.13%) |
|  | XBB.1.5.32 |  | 2 (0.09%) |
|  | XBB.1.5.33 |  | 2 (0.09%) |
|  | XBB.1.5.5 |  | 2 (0.09%) |
|  | XBB.1.5.8 |  | 2 (0.09%) |
|  | XBB.1.7 |  | 2 (0.09%) |
|  | XBB.1.9.4 |  | 2 (0.09%) |
|  | XBB.2.3.1 |  | 2 (0.09%) |
|  | XBB.5 |  | 2 (0.09%) |
|  | XBB.6 |  | 2 (0.09%) |
|  | XBB.1.1 |  | 1 (0.04%) |
|  | XBB.1.12 |  | 1 (0.04%) |
|  | XBB.1.17.1 |  | 1 (0.04%) |
|  | XBB.1.19 |  | 1 (0.04%) |
|  | XBB.1.22.1 |  | 1 (0.04%) |
|  | XBB.1.22.2 |  | 1 (0.04%) |
|  | XBB.1.27 |  | 1 (0.04%) |
|  | XBB.1.4 |  | 1 (0.04%) |
|  | XBB.1.5.16 |  | 1 (0.04%) |
|  | XBB.1.5.20 |  | 1 (0.04%) |
|  | XBB.1.5.23 |  | 1 (0.04%) |
|  | XBB.1.5.31 |  | 1 (0.04%) |
|  | XBB.1.5.39 |  | 1 (0.04%) |
|  | XBB.1.5.4 |  | 1 (0.04%) |
|  | XBB.1.9.5 |  | 1 (0.04%) |
|  | XBB.2.1 |  | 1 (0.04%) |
|  | XBB.2.7.1 |  | 1 (0.04%) |
|  | XBB.2.8 |  | 1 (0.04%) |
|  | XBB.3.2 |  | 1 (0.04%) |
|  | FD.1 |  | 1 (0.04%) |
| BQ.1* | BQ.1.1 | 182<br>(6.37%) | 48 (26.37%) |
|  | BQ.1 |  | 25 (13.74%) |
|  | BQ.1.2 |  | 6 (3.30%) |
|  | BQ.1.22 |  | 6 (3.30%) |
|  | BQ.1.23 |  | 6 (3.30%) |
|  | BQ.1.1.3 |  | 5 (2.75%) |
|  | BQ.1.1.51 |  | 5 (2.75%) |
|  | BQ.1.2.1 |  | 5 (2.75%) |
|  | BQ.1.1.18 |  | 4 (2.20%) |
|  | BQ.1.1.8 |  | 4 (2.20%) |
|  | BQ.1.10 |  | 4 (2.20%) |
|  | BQ.1.12 |  | 4 (2.20%) |
|  | BQ.1.13.1 |  | 4 (2.20%) |
|  | BQ.1.1.22 |  | 3 (1.65%) |
|  | BQ.1.1.32 |  | 3 (1.65%) |
|  | BQ.1.1.45 |  | 3 (1.65%) |
|  | BQ.1.15 |  | 3 (1.65%) |
|  | BQ.1.5 |  | 3 (1.65%) |
|  | BQ.1.9 |  | 3 (1.65%) |
|  | BQ.1.1.4 |  | 2 (1.10%) |
|  | BQ.1.1.5 |  | 2 (1.10%) |
|  | BQ.1.1.7 |  | 2 (1.10%) |
|  | BQ.1.8 |  | 2 (1.10%) |
|  | DP.1 |  | 2 (1.10%) |
|  | BQ.1.1.11 |  | 1 (0.55%) |
|  | BQ.1.1.12 |  | 1 (0.55%) |
|  | BQ.1.1.24 |  | 1 (0.55%) |
|  | BQ.1.1.31 |  | 1 (0.55%) |
|  | BQ.1.1.35 |  | 1 (0.55%) |
|  | BQ.1.1.38 |  | 1 (0.55%) |
|  | BQ.1.1.41 |  | 1 (0.55%) |
|  | BQ.1.1.47 |  | 1 (0.55%) |
|  | BQ.1.1.52 |  | 1 (0.55%) |
|  | BQ.1.1.59 |  | 1 (0.55%) |
|  | BQ.1.1.67 |  | 1 (0.55%) |
|  | BQ.1.1.68 |  | 1 (0.55%) |
|  | BQ.1.1.74 |  | 1 (0.55%) |
|  | BQ.1.10.3 |  | 1 (0.55%) |
|  | BQ.1.18 |  | 1 (0.55%) |
|  | BQ.1.24 |  | 1 (0.55%) |
|  | BQ.1.25 |  | 1 (0.55%) |
|  | BQ.1.25.1 |  | 1 (0.55%) |
|  | BQ.1.3 |  | 1 (0.55%) |
|  | BQ.1.32 |  | 1 (0.55%) |
|  | BQ.1.8.1 |  | 1 (0.55%) |
|  | BQ.1.8.2 |  | 1 (0.55%) |
|  | DU.1 |  | 1 (0.55%) |
|  | EE.2 |  | 1 (0.55%) |
|  | EF.1 |  | 1 (0.55%) |
|  | EF.1.1.1 |  | 1 (0.55%) |
|  | EW.2 |  | 1 (0.55%) |
|  | FC.1 |  | 1 (0.55%) |
| BA.2.75* | CH.1.1 | 157<br>(5.50%) | 16 (10.19%) |
|  | BN.1.2 |  | 12 (7.64%) |
|  | BN.1 |  | 9 (5.73%) |
|  | BY.1 |  | 9 (5.73%) |
|  | CH.1.1.1 |  | 9 (5.73%) |
|  | BR.2.1 |  | 8 (5.10%) |
|  | CH.1.1.3 |  | 7 (4.46%) |
|  | BN.1.4.5 |  | 6 (3.82%) |
|  | CH.1.1.18 |  | 6 (3.82%) |
|  | BN.1.3.1 |  | 5 (3.18%) |
|  | BN.1.4 |  | 5 (3.18%) |
|  | BA.2.75 |  | 5 (3.18%) |
|  | BA.2.75.2 |  | 4 (2.55%) |
|  | BL.1 |  | 4 (2.55%) |
|  | BN.1.1 |  | 4 (2.55%) |
|  | BN.1.1.1 |  | 4 (2.55%) |
|  | BN.1.3 |  | 4 (2.55%) |
|  | BA.2.75.6 |  | 3 (1.91%) |
|  | BM.1.1.3 |  | 3 (1.91%) |
|  | CH.1.1.11 |  | 3 (1.91%) |
|  | CH.1.1.7 |  | 3 (1.91%) |
|  | BA.2.75.1 |  | 2 (1.27%) |
|  | BL.2 |  | 2 (1.27%) |
|  | BN.1.3.5 |  | 2 (1.27%) |
|  | BN.1.3.7 |  | 2 (1.27%) |
|  | BN.2.1 |  | 2 (1.27%) |
|  | BR.2 |  | 2 (1.27%) |
|  | BA.2.75.10 |  | 1 (0.64%) |
|  | BA.2.75.4 |  | 1 (0.64%) |
|  | BL.1.3 |  | 1 (0.64%) |
|  | BM.1.1 |  | 1 (0.64%) |
|  | BM.4.1.1 |  | 1 (0.64%) |
|  | BN.1.11 |  | 1 (0.64%) |
|  | BN.1.2.2 |  | 1 (0.64%) |
|  | BN.1.2.3 |  | 1 (0.64%) |
|  | BN.1.2.4 |  | 1 (0.64%) |
|  | BN.1.3.8 |  | 1 (0.64%) |
|  | BN.1.5 |  | 1 (0.64%) |
|  | BN.5 |  | 1 (0.64%) |
|  | BY.1.1 |  | 1 (0.64%) |
|  | CA.3 |  | 1 (0.64%) |
|  | DV.2 |  | 1 (0.64%) |
|  | EP.1 |  | 1 (0.64%) |
| BA.5* | BA.5 | 74<br>(2.59%) | 9 (12.16%) |
|  | BA.5.2 |  | 5 (6.76%) |
|  | BF.7.14 |  | 5 (6.76%) |
|  | BA.5.2.48 |  | 4 (5.41%) |
|  | CK.1 |  | 4 (5.41%) |
|  | DY.2 |  | 4 (5.41%) |
|  | DY.4 |  | 4 (5.41%) |
|  | BA.5.3 |  | 3 (4.05%) |
|  | BE.9 |  | 3 (4.05%) |
|  | CK.3 |  | 3 (4.05%) |
|  | BA.5.1 |  | 2 (2.70%) |
|  | BA.5.2.1 |  | 2 (2.70%) |
|  | BE.1.1.1 |  | 2 (2.70%) |
|  | BF.5 |  | 2 (2.70%) |
|  | BF.7.15 |  | 2 (2.70%) |
|  | BF.7.6 |  | 2 (2.70%) |
|  | BA.5.1.28 |  | 1 (1.35%) |
|  | BA.5.1.36 |  | 1 (1.35%) |
|  | BA.5.2.26 |  | 1 (1.35%) |
|  | BA.5.2.34 |  | 1 (1.35%) |
|  | BA.5.2.49 |  | 1 (1.35%) |
|  | BA.5.2.9 |  | 1 (1.35%) |
|  | BA.5.3.1 |  | 1 (1.35%) |
|  | BE.1.2 |  | 1 (1.35%) |
|  | BF.26 |  | 1 (1.35%) |
|  | BF.7 |  | 1 (1.35%) |
|  | BF.7.4 |  | 1 (1.35%) |
|  | BF.7.4.1 |  | 1 (1.35%) |
|  | BF.7.5.1 |  | 1 (1.35%) |
|  | BW.1.1 |  | 1 (1.35%) |
|  | CG.1 |  | 1 (1.35%) |
|  | CL.1 |  | 1 (1.35%) |
|  | CP.7 |  | 1 (1.35%) |
|  | CQ.2 |  | 1 (1.35%) |
| Other Lineages | B.1.14 | 68<br>(2.38%) | 23 (33.82%) |
|  | B.1.617.2 |  | 14 (20.59%) |
|  | B.1.1 |  | 10 (14.71%) |
|  | B |  | 7 (10.29%) |
|  | B.1.1.161 |  | 6 (8.82%) |
|  | B.1 |  | 5 (7.35%) |
|  | B.26 |  | 1 (1.47%) |
|  | B.55 |  | 1 (1.47%) |
|  | B.6 |  | 1 (1.47%) |
| Other Omicron Lineages | BA.2 | 60<br>(2.10%) | 29 (48.33%) |
|  | B.1.1.529 |  | 6 (10%) |
|  | BA.2.10 |  | 6 (10%) |
|  | BA.2.23 |  | 6 (10%) |
|  | BA.2.76 |  | 3 (5%) |
|  | CM.12 |  | 3 (5%) |
|  | BA.2.10.1 |  | 2 (3.33%) |
|  | BA.2.12.1 |  | 1 (1.67%) |
|  | BA.2.3.20 |  | 1 (1.67%) |
|  | BA.2.31 |  | 1 (1.67%) |
|  | BA.2.74 |  | 1 (1.67%) |
|  | BA.4.6 |  | 1 (1.67%) |
| Other Recombinant Lineages | XBF | 34<br>(1.19%) | 10 (29.41%) |
|  | XAP |  | 8 (23.53%) |
|  | XBH |  | 4 (11.76%) |
|  | XAR |  | 3 (8.82%) |
|  | XBL |  | 2 (5.88%) |
|  | XT |  | 2 (5.88%) |
|  | XAH |  | 1 (2.94%) |
|  | XAY.2 |  | 1 (2.94%) |
|  | XBF.5 |  | 1 (2.94%) |
|  | XBN |  | 1 (2.94%) |
|  | XM |  | 1 (2.94%) |
| Grand Total |  | 2856 (100%) |  |

***Figure 1*** shows the evolutionary relationship of the XBB.1.16* lineage (clade 22F) detected in our dataset with other lineages. Prior to the fifth week of 2023, the sequences from India on GISAID were dominated by the BQ.1* lineage (17.07%), followed by the XBB.1* (14.24%) and XBB.2* (12.73%) (***Figure 2***). It is important to emphasize that even though India was reporting a minimal number of COVID-19 cases during this period, the Government of India issued an advisory to screen international travellers due to a rise in COVID-19 cases in countries like China, Singapore, Hong Kong, Republic of Korea, Thailand and Japan. Therefore, the primary focus of SARS-CoV-2 sequencing activity during this period in India was on international travellers [15]. The XBB.1.16* lineage first appeared in Indian sequences on 25^th^ December 2022 in a sample collected in Chennai, Tamil Nadu. Since the fifth week of 2023, the XBB.1.16* lineage has grown from 9.30% to 79.17% in the 13^th^ week of 2023 (***Figure 3***).

**Figure 1:**
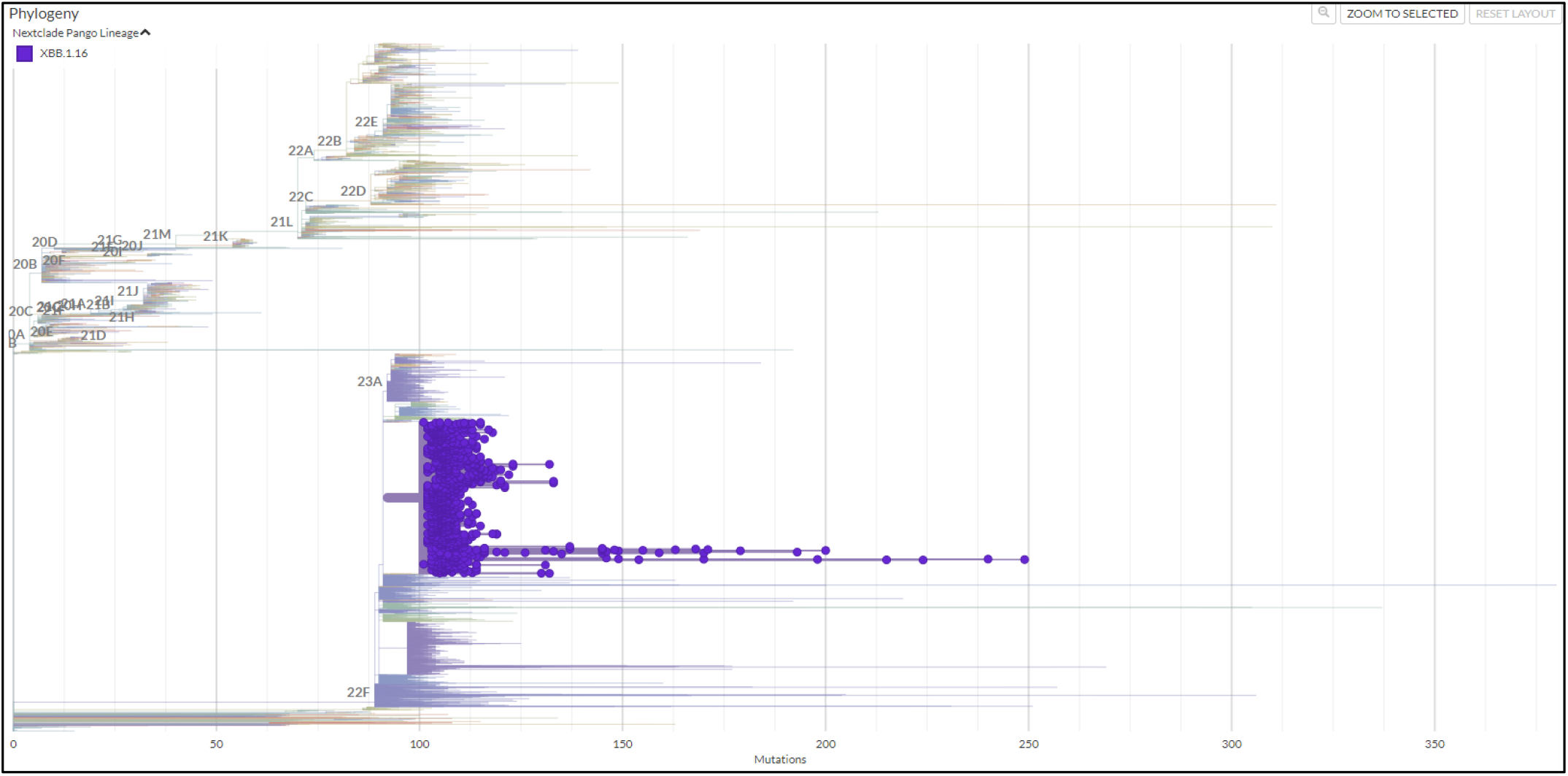
Evolutionary relationship of XBB.1.16* lineage (Clade 22F) with other clades in India (Data from GISAID between 1^st^ December 2022 and 8^th^ April 2023)

**Figure 2:**
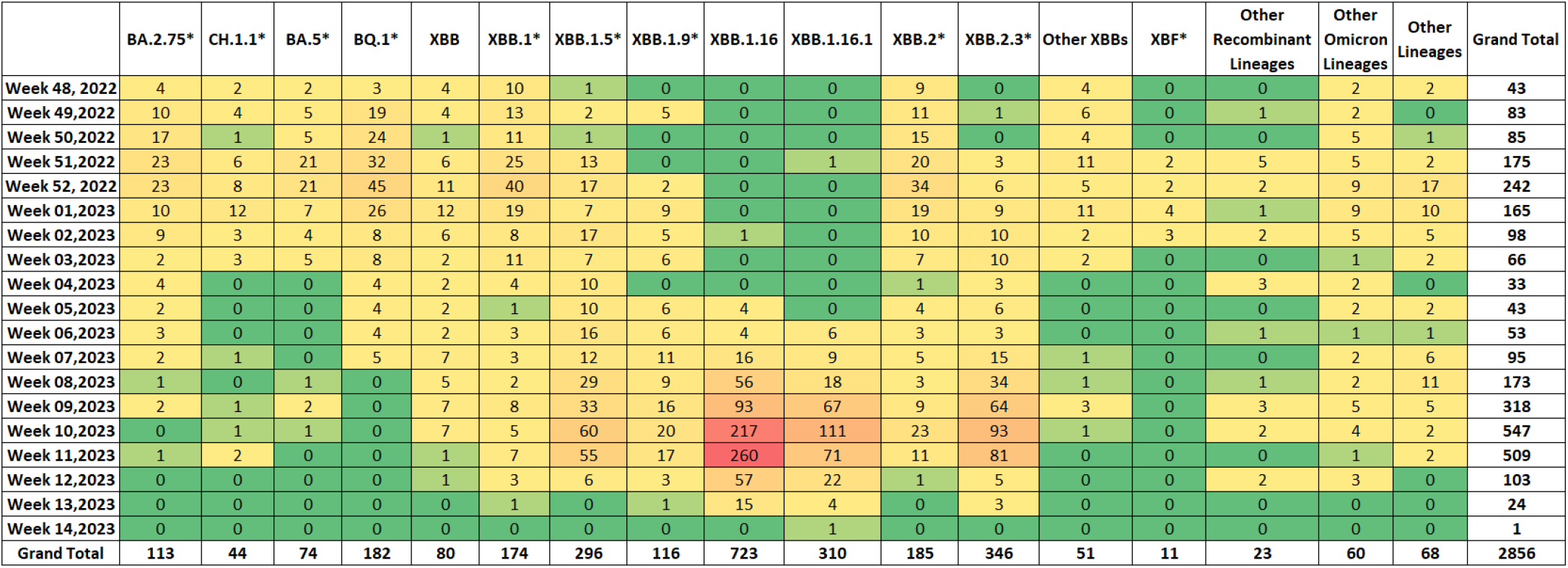
Heatmap showing the distribution of SARS-CoV-2 lineages in India (Data from GISAID between 1^st^ December 2022 and 8^th^ April 2023)

**Figure 3:**
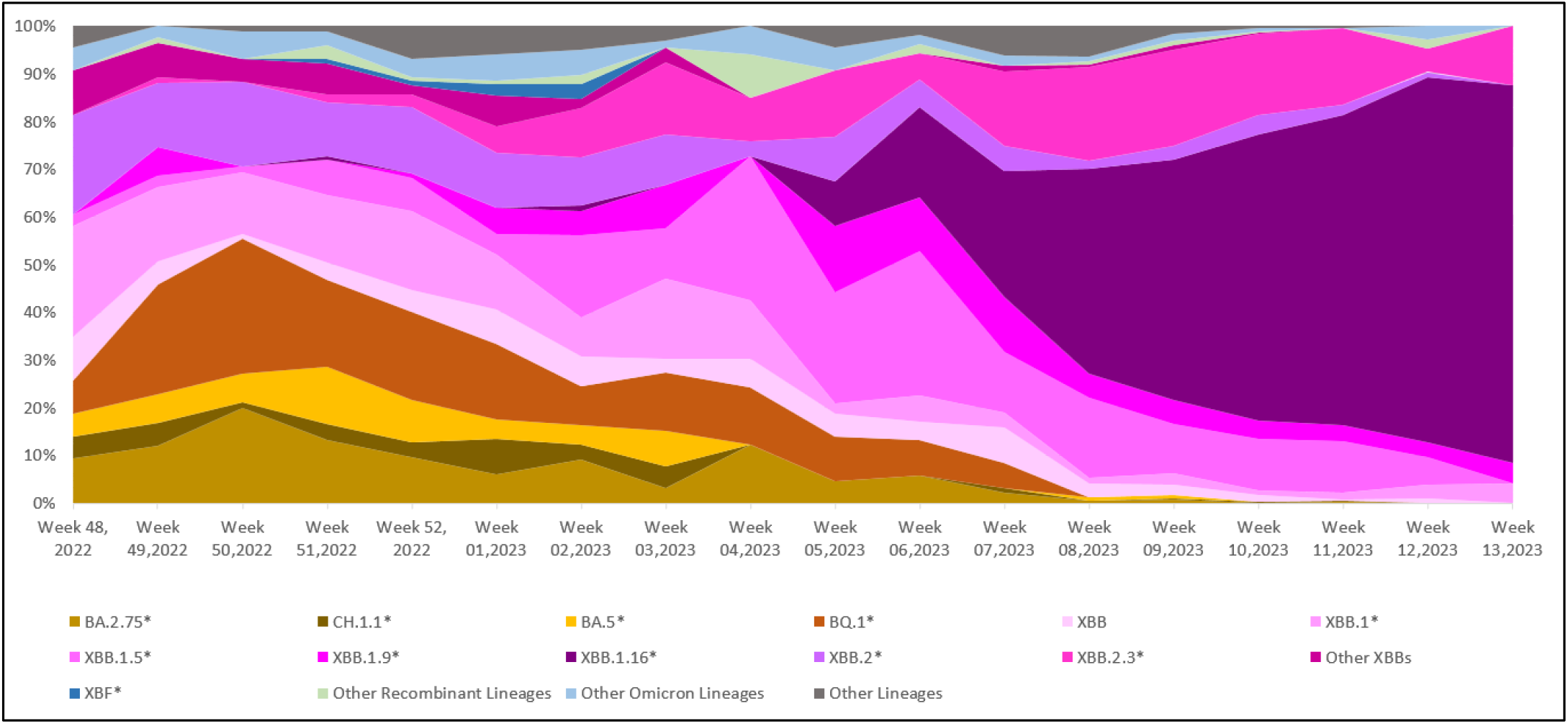
Temporal distribution of XBB.1.16* and other Omicron lineages in circulation in India (Data from GISAID between 1^st^ December 2022 and 8^th^ April 2023)

The recent increase in COVID-19 cases in India appeared to be linked to the emergence of the XBB.1.16* lineage, as its prevalence has risen concurrently with the upsurge in cases. ***Figure 4*** shows the distribution of XBB.1.16* in Indian states, with the highest number of sequences from Maharashtra (42.40%) and Gujarat (37.27%) deposited on the GISAID database.

**Figure 4:**
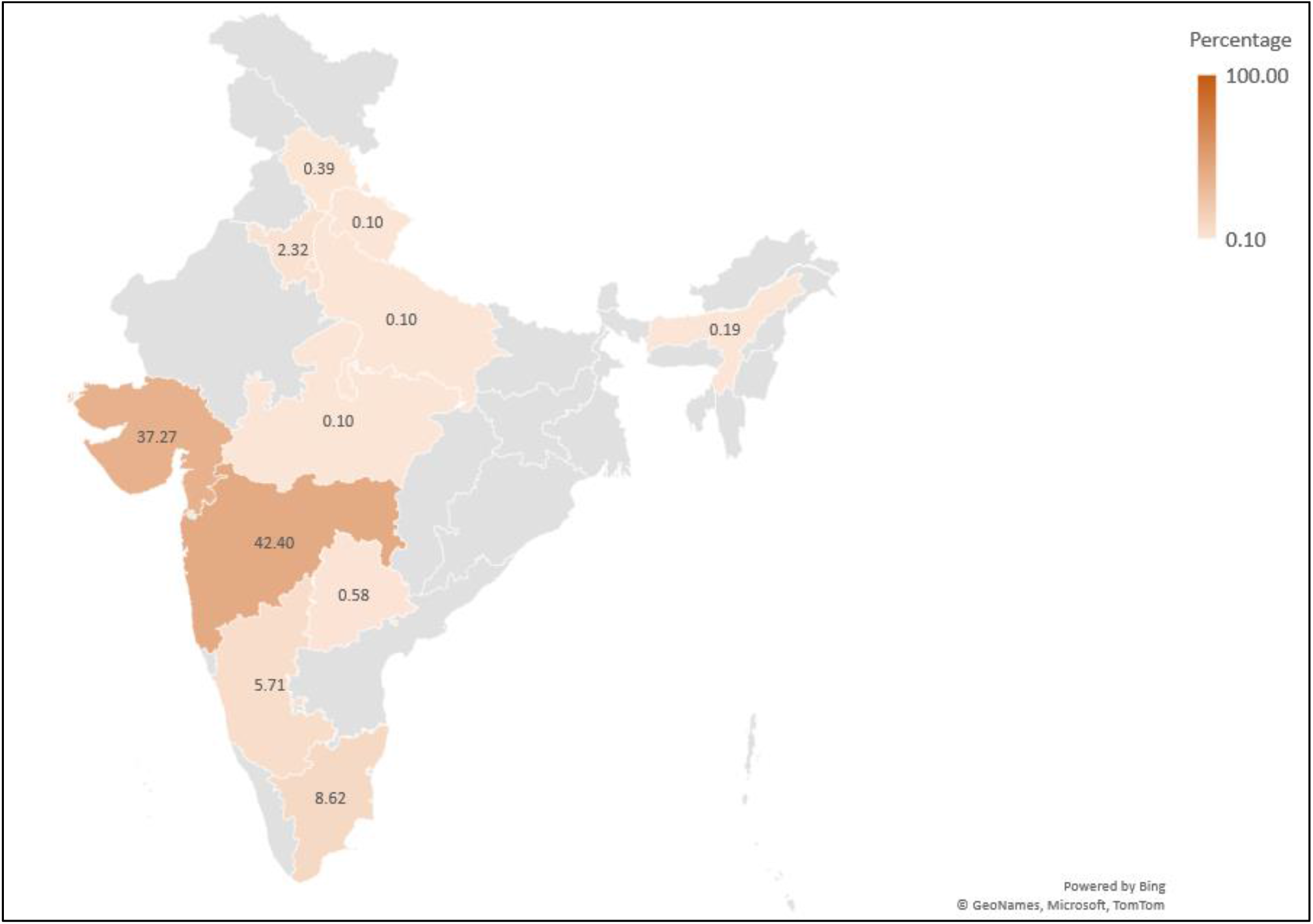
Distribution of XBB.1.16* lineage in India (Data from GISAID between 1^st^ December 2022 and 8^th^ April 2023)

### 3.2. Distribution of SARS-CoV-2 Lineages in Maharashtra

The lineage distribution of SARS-CoV-2 in Maharashtra showed a pattern consistent with that observed throughout India (***Figure 5 and 6***) with XBB.1.16* increasing from 3.85% in the second week of 2023 to 79.17% in the 13^th^ week of 2023. In Maharashtra, XBB.1.16* was first found on 11^th^ January 2023 in a sample collected in Mumbai, Maharashtra. ***Figure 7*** shows the distribution of XBB.1.16* in various districts of Maharashtra, with most sequences from the Pune district (60.27%) followed by Mumbai suburban (9.13%) and Aurangabad and Amravati (6.16%, each).

**Figure 5:**
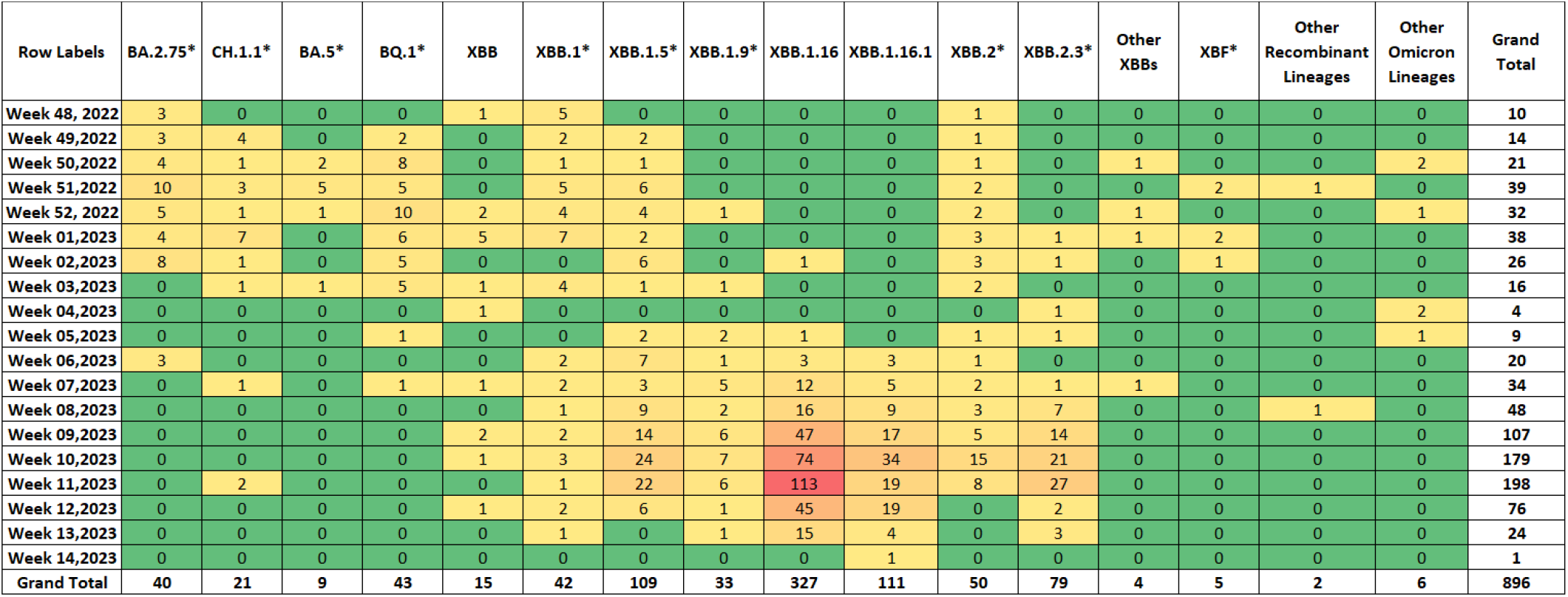
Heatmap showing the distribution of SARS-CoV-2 lineages in Maharashtra (Data from GISAID between 1^st^ December 2022 and 8^th^ April 2023)

**Figure 6:**
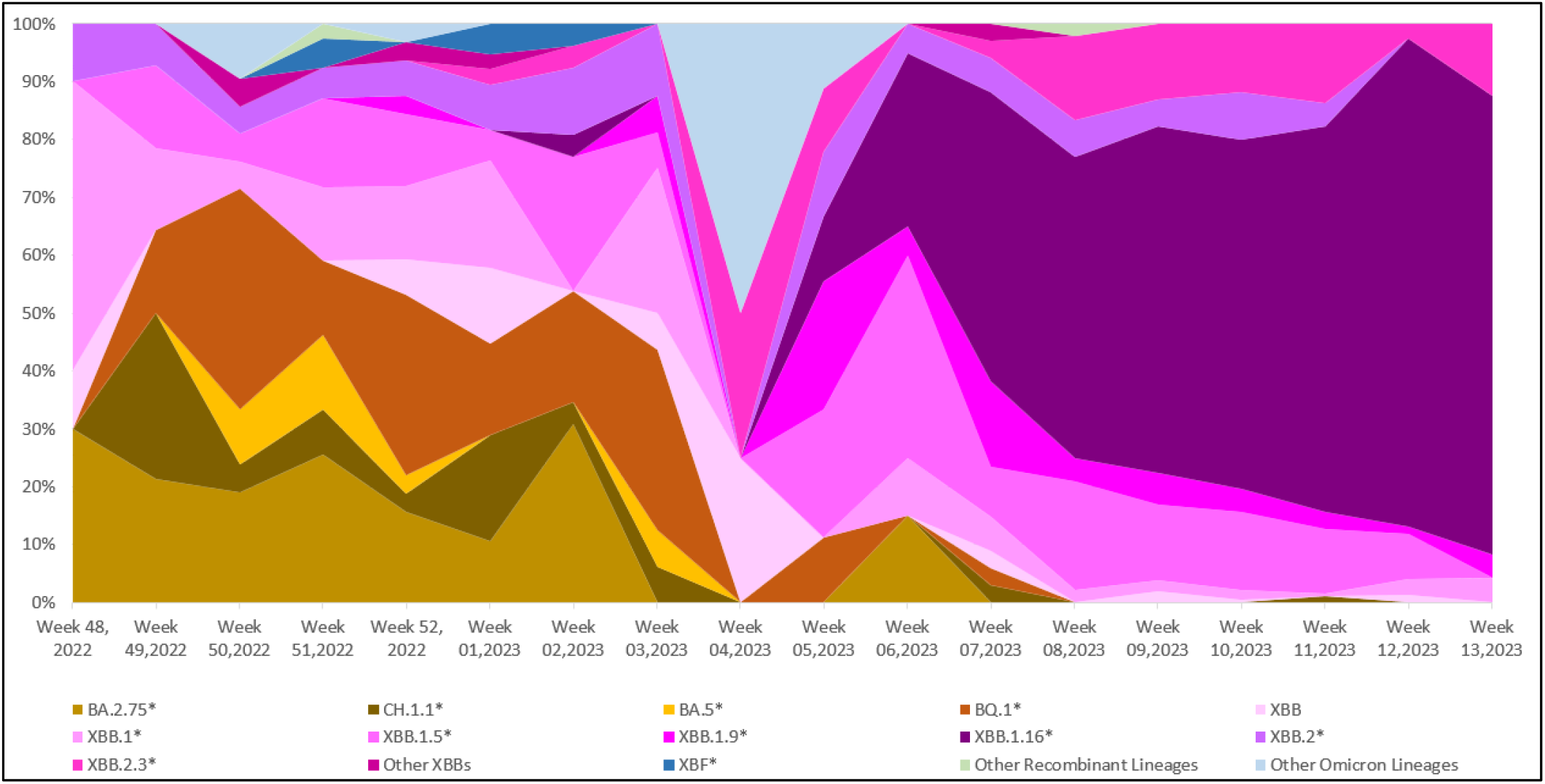
Temporal distribution of XBB.1.16* and other Omicron lineages in circulation in Maharashtra (Data from GISAID between 1^st^ December 2022 and 8^th^ April 2023)

**Figure 7:**
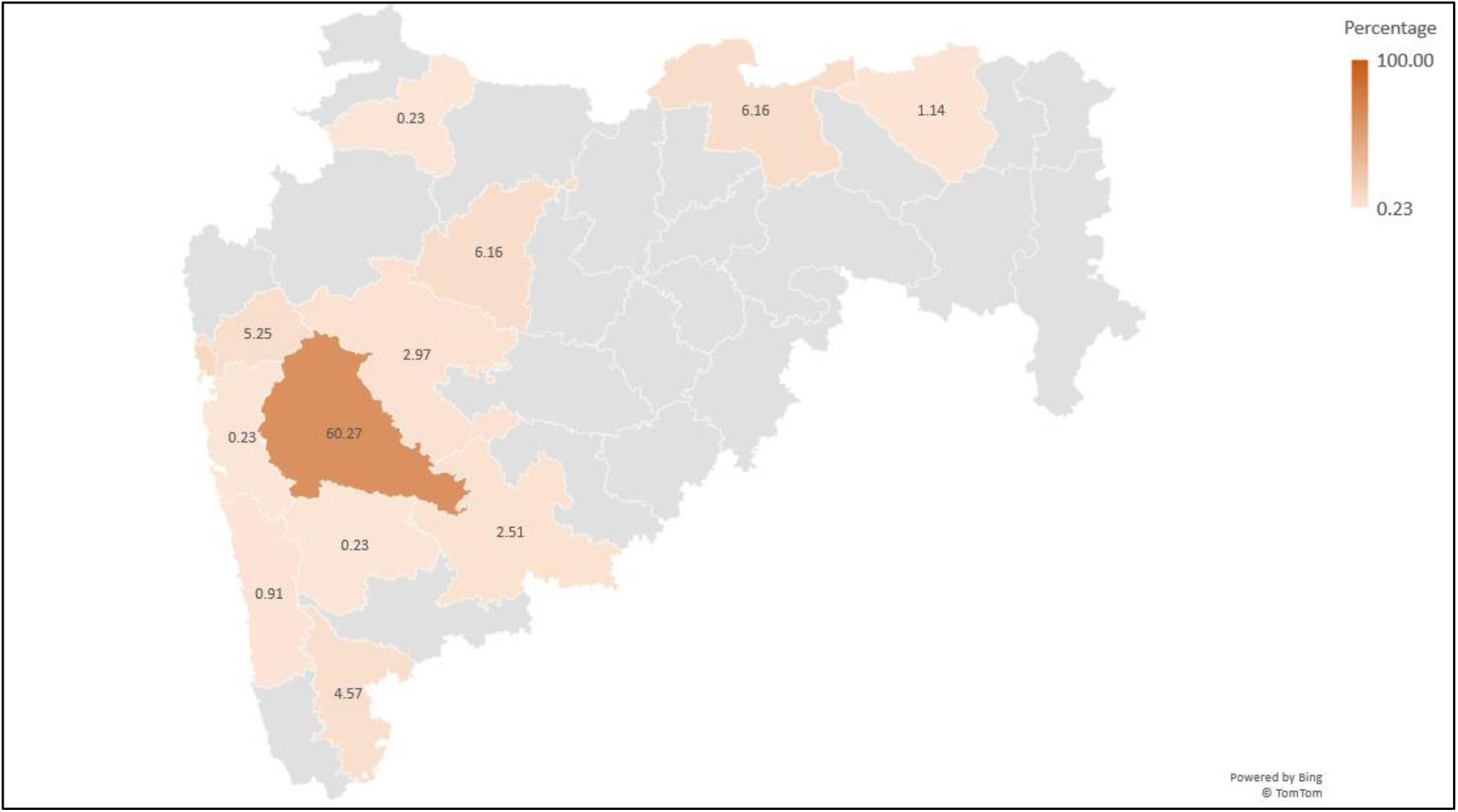
Distribution of XBB.1.16* lineage in Maharashtra (Data from GISAID between 1^st^ December 2022 and 8^th^ April 2023)

### 3.3. Comparison of Demographic and Clinical Characteristics of XBB.1.16* lineage and Other Omicron Lineages in Maharashtra

A total of 693 RT-PCR-positive cases in Maharashtra were included in the demographic study. ***Table 2*** shows the lineage distribution of the 693 cases, of which XBB.1.16* is the dominant SARS-CoV-2 lineage (70.56%), followed by other Omicron lineages (29.29%).

**Table 2:**
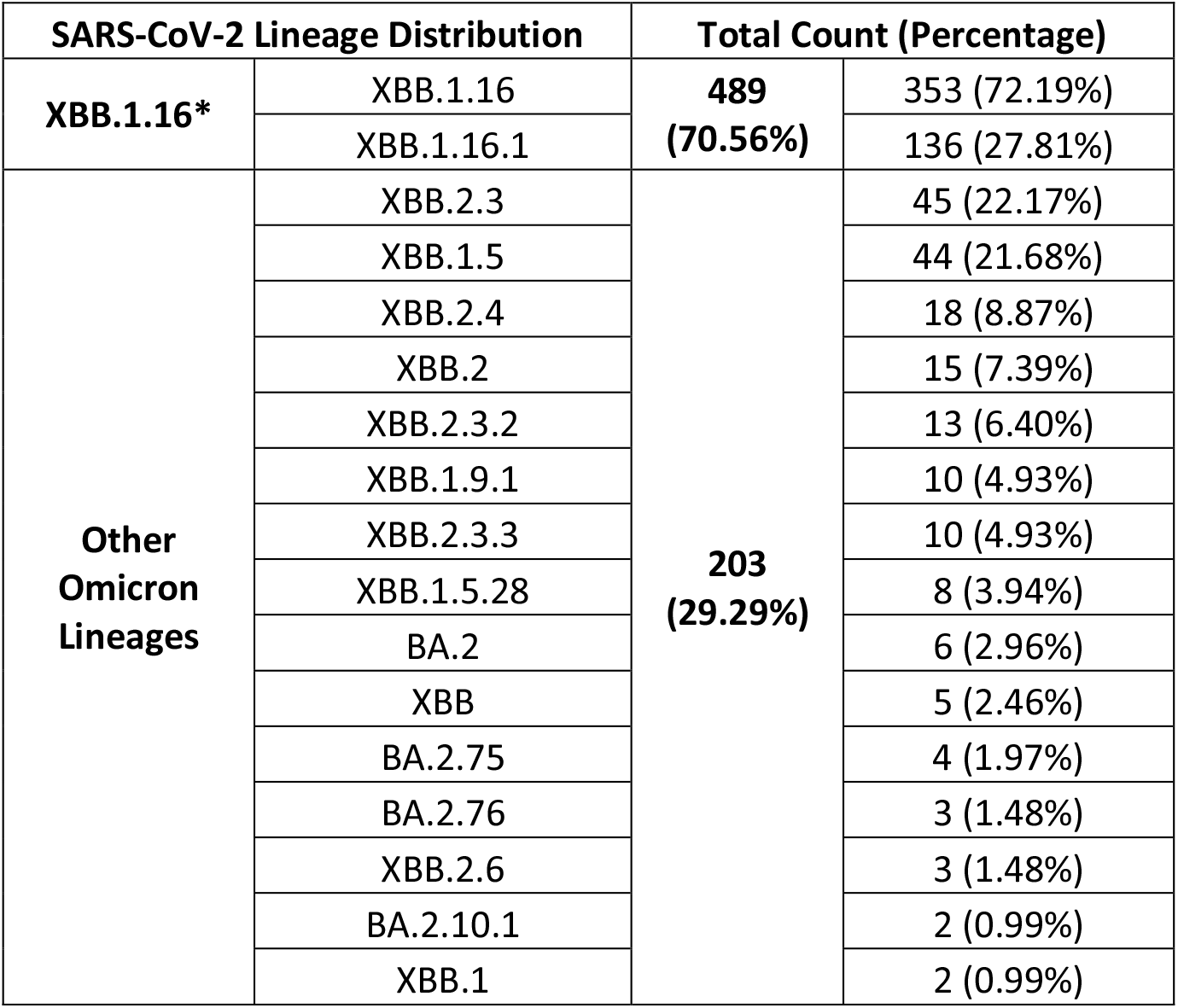
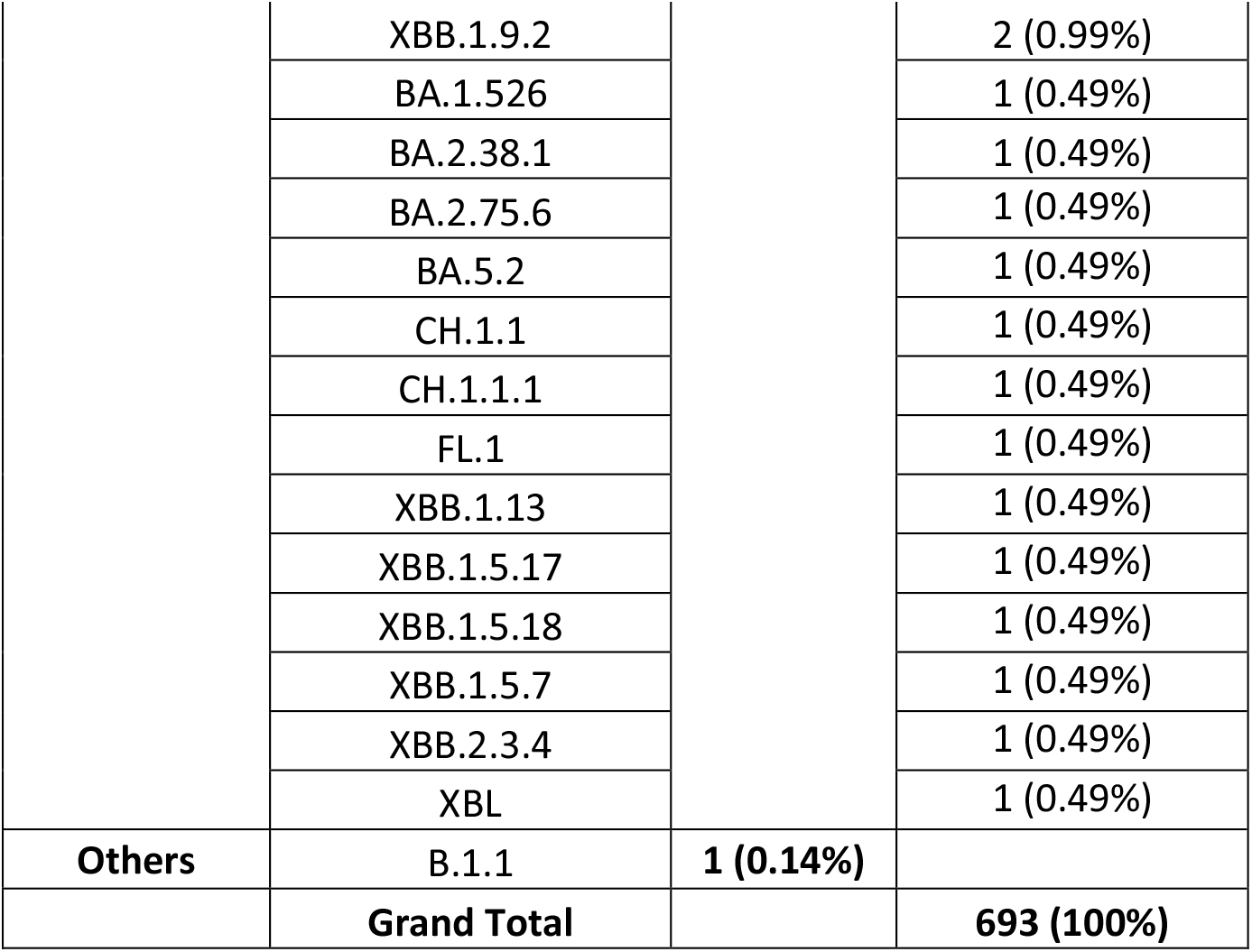
SARS-CoV-2 lineages identified among 693 RT-PCR-positive cases in Maharashtra

Of the 693 RT-PCR-positive cases, 52.2% were males, and 47.8% were females (Male: Female :: 1.09:1). The median age of cases infected with XBB.1.16* was 35.0 (IQR: 24.0 – 52.5) years, and for other Omicron lineages it was 35.5 (IQR: 25.0 – 53.0) years. The age group 20 years to 39 years was predominantly affected. However, there was no statistically significant difference in the age-wise distribution of cases among XBB.1.16* and other lineages (***Table 3***).

**Table 3:** Demographic characteristics of XBB.1.16* and other Omicron lineages in Maharashtra

| Demographic Characteristics | XBB.1.16* |  | Other Lineages | Grand Total | p-value |
| --- | --- | --- | --- | --- | --- |
|  | XBB.1.16 | XBB.1.16.1 |  |  |  |
| 1. Gender-wise distribution |  |  |  |  | 0.111 |
| Male | 191 (54.1%) | 74 (54.4%) | 97 (47.5%) | 362 (52.2%) |  |
| Female | 162 (45.9%) | 62 (45.6%) | 107 (52.5%) | 331 (47.8%) |  |
| 2. Median Age |  |  |  |  | 0.883 |
|  | 33.0<br>(IQR: 22.0 – 54.0) | 39.0<br>(IQR: 29.0 – 50.0) | 35.5<br>(IQR: 25.0 – 53.0) |  |  |
| 3. Age-wise distribution (Age in years) |  |  |  |  | 0.772 |
| 0 to 9 | 24 (6.8%) | 6 (4.4%) | 14 (6.9%) | 44 (6.4%) |  |
| 10 to 19 | 41 (11.6%) | 5 (3.7%) | 17 (8.3%) | 63 (9.1%) |  |
| 20 to 29 | 75 (21.3%) | 24 (17.6%) | 46 (22.5%) | 145 (20.9%) |  |
| 30 to 39 | 71 (20.1%) | 34 (25.0%) | 36 (17.6%) | 141 (20.3%) |  |
| 40 to 49 | 40 (11.3%) | 32 (23.5%) | 25 (12.3%) | 97 (14.0%) |  |
| 50 to 59 | 33 (9.4%) | 13 (9.6%) | 23 (11.3%) | 69 (10.0%) |  |
| 60 and above | 69 (19.5%) | 22 (16.2%) | 43 (21.1%) | 134 (19.3%) |  |
| 4. Area-wise distribution |  |  |  |  | - |
| Ahmednagar | 19 (5.4%) | 0 | 5 (2.5%) | 24 (3.5%) |  |
| Akola | 3 (0.8%) | 0 | 8 (3.9%) | 11 (1.6%) |  |
| Amravati | 18 (5.1%) | 6 (4.4%) | 19 (9.3%) | 43 (6.2%) |  |
| Aurangabad | 26 (7.4%) | 1 (0.7%) | 11 (5.4%) | 38 (5.5%) |  |
| Beed | 0 | 1 (0.7%) | 0 | 1 (0.1%) |  |
| Kolhapur | 13 (3.7%) | 9 (6.6%) | 2 (1.0%) | 24 (3.5%) |  |
| Mumbai | 13 (3.7%) | 14 (10.3%) | 27 (13.2%) | 54 (7.8%) |  |
| Nagpur | 06 (1.7%) | 0 | 12 (5.9%) | 18 (2.6%) |  |
| Pune | 216 (61.2%) | 96 (70.6%) | 105 (51.5%) | 417 (60.2%) |  |
| Raigad | 0 | 1 (0.7%) | 1 (0.5%) | 2 (0.3%) |  |
| Ratnagiri | 3 (0.8%) | 1 (0.7%) | 3 (1.5%) | 7 (1.0%) |  |
| Satara | 1 (0.3%) | 0 | 0 | 1 (0.1%) |  |
| Solapur | 17 (4.8%) | 2 (1.5%) | 1 (0.5%) | 20 (2.9%) |  |
| Thane | 18 (5.1%) | 5 (3.7%) | 10 (4.9%) | 33 (4.8%) |  |

Of the 693 cases included in the study, 386 (55.70%) consented to share their clinical details. ***Table 4*** summarizes the clinical characteristics, vaccination status and the outcome of these 386 cases. Of the 386 cases, 276 (71.50%) were XBB.1.16*. Most cases of XBB.1.16* had the symptomatic disease (92%) with mild symptoms, with fever (67%) being the most common symptom. Out of 276 XBB.1.16* cases, comorbidity was reported in 17.7% of cases, of which hypertension was the most common condition reported (47.9%), followed by diabetes mellitus (39.6%) and asthma (12.5%). There was no statistically significant difference in the presence or absence of symptoms, history of previous COVID-19 infection, underlying comorbid conditions or clinical manifestations between individuals infected with XBB.1.16* and those infected with other Omicron lineages in this study.

**Table 4:**
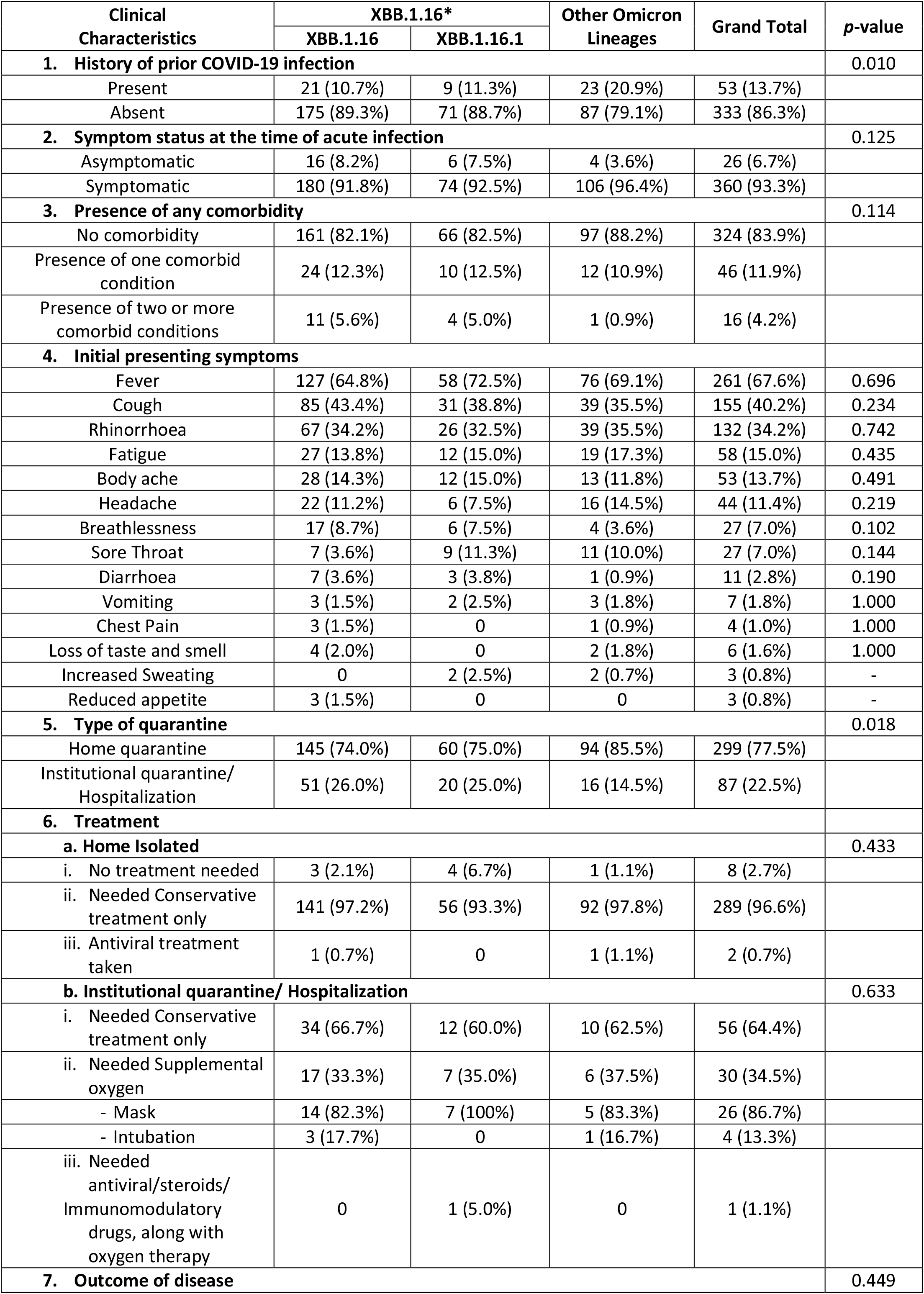
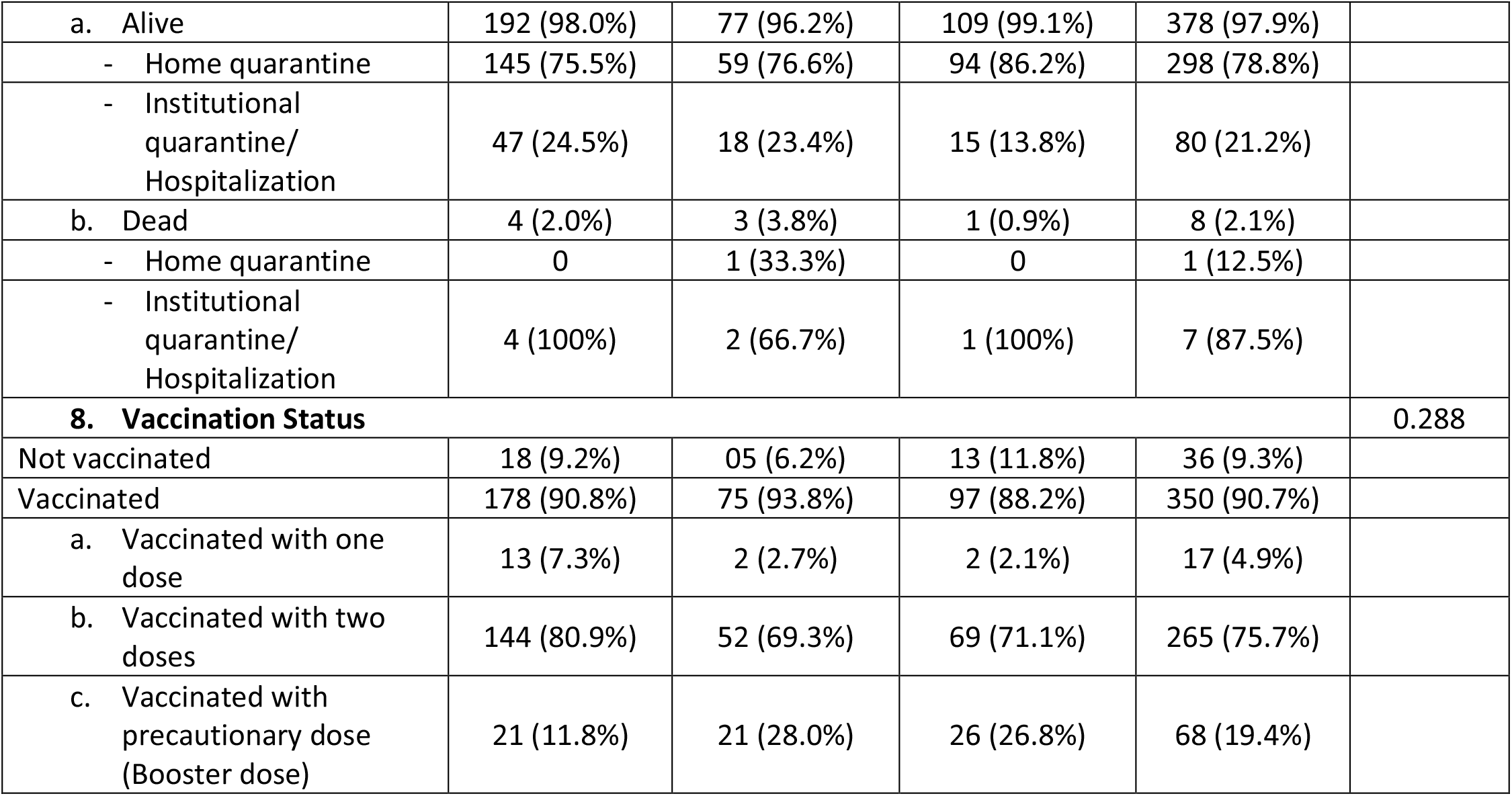
Clinical characteristics of XBB.1.16* and other Omicron lineages in Maharashtra

| Clinical Characteristics | XBB.1.16* |  | Other Omicron Lineages | Grand Total | p-value |
| --- | --- | --- | --- | --- | --- |
|  | XBB.1.16 | XBB.1.16.1 |  |  |  |
| 1. History of prior COVID-19 infection |  |  |  |  | 0.010 |
| Present | 21 (10.7%) | 9 (11.3%) | 23 (20.9%) | 53 (13.7%) |  |
| Absent | 175 (89.3%) | 71 (88.7%) | 87 (79.1%) | 333 (86.3%) |  |
| 2. Symptom status at the time of acute infection |  |  |  |  | 0.125 |
| Asymptomatic | 16 (8.2%) | 6 (7.5%) | 4 (3.6%) | 26 (6.7%) |  |
| Symptomatic | 180 (91.8%) | 74 (92.5%) | 106 (96.4%) | 360 (93.3%) |  |
| 3. Presence of any comorbidity |  |  |  |  | 0.114 |
| No comorbidity | 161 (82.1%) | 66 (82.5%) | 97 (88.2%) | 324 (83.9%) |  |
| Presence of one comorbid condition | 24 (12.3%) | 10 (12.5%) | 12 (10.9%) | 46 (11.9%) |  |
| Presence of two or more comorbid conditions | 11 (5.6%) | 4 (5.0%) | 1 (0.9%) | 16 (4.2%) |  |
| 4. Initial presenting symptoms |  |  |  |  |  |
| Fever | 127 (64.8%) | 58 (72.5%) | 76 (69.1%) | 261 (67.6%) | 0.696 |
| Cough | 85 (43.4%) | 31 (38.8%) | 39 (35.5%) | 155 (40.2%) | 0.234 |
| Rhinorrhoea | 67 (34.2%) | 26 (32.5%) | 39 (35.5%) | 132 (34.2%) | 0.742 |
| Fatigue | 27 (13.8%) | 12 (15.0%) | 19 (17.3%) | 58 (15.0%) | 0.435 |
| Body ache | 28 (14.3%) | 12 (15.0%) | 13 (11.8%) | 53 (13.7%) | 0.491 |
| Headache | 22 (11.2%) | 6 (7.5%) | 16 (14.5%) | 44 (11.4%) | 0.219 |
| Breathlessness | 17 (8.7%) | 6 (7.5%) | 4 (3.6%) | 27 (7.0%) | 0.102 |
| Sore Throat | 7 (3.6%) | 9 (11.3%) | 11 (10.0%) | 27 (7.0%) | 0.144 |
| Diarrhoea | 7 (3.6%) | 3 (3.8%) | 1 (0.9%) | 11 (2.8%) | 0.190 |
| Vomiting | 3 (1.5%) | 2 (2.5%) | 3 (1.8%) | 7 (1.8%) | 1.000 |
| Chest Pain | 3 (1.5%) | 0 | 1 (0.9%) | 4 (1.0%) | 1.000 |
| Loss of taste and smell | 4 (2.0%) | 0 | 2 (1.8%) | 6 (1.6%) | 1.000 |
| Increased Sweating | 0 | 2 (2.5%) | 2 (0.7%) | 3 (0.8%) | - |
| Reduced appetite | 3 (1.5%) | 0 | 0 | 3 (0.8%) | - |
| 5. Type of quarantine |  |  |  |  | 0.018 |
| Home quarantine | 145 (74.0%) | 60 (75.0%) | 94 (85.5%) | 299 (77.5%) |  |
| Institutional quarantine/<br>Hospitalization | 51 (26.0%) | 20 (25.0%) | 16 (14.5%) | 87 (22.5%) |  |
| 6. Treatment |  |  |  |  |  |
| a. Home Isolated |  |  |  |  | 0.433 |
| i. No treatment needed | 3 (2.1%) | 4 (6.7%) | 1 (1.1%) | 8 (2.7%) |  |
| ii. Needed Conservative treatment only | 141 (97.2%) | 56 (93.3%) | 92 (97.8%) | 289 (96.6%) |  |
| iii. Antiviral treatment taken | 1 (0.7%) | 0 | 1 (1.1%) | 2 (0.7%) |  |
| b. Institutional quarantine/ Hospitalization |  |  |  |  | 0.633 |
| i. Needed Conservative treatment only | 34 (66.7%) | 12 (60.0%) | 10 (62.5%) | 56 (64.4%) |  |
| ii. Needed Supplemental oxygen | 17 (33.3%) | 7 (35.0%) | 6 (37.5%) | 30 (34.5%) |  |
| - Mask | 14 (82.3%) | 7 (100%) | 5 (83.3%) | 26 (86.7%) |  |
| - Intubation | 3 (17.7%) | 0 | 1 (16.7%) | 4 (13.3%) |  |
| iii. Needed antiviral/steroids/<br>Immunomodulatory drugs, along with oxygen therapy | 0 | 1 (5.0%) | 0 | 1 (1.1%) |  |
| 7. Outcome of disease |  |  |  |  | 0.449 |
| a. Alive | 192 (98.0%) | 77 (96.2%) | 109 (99.1%) | 378 (97.9%) |  |
| - Home quarantine | 145 (75.5%) | 59 (76.6%) | 94 (86.2%) | 298 (78.8%) |  |
| - Institutional quarantine/<br>Hospitalization | 47 (24.5%) | 18 (23.4%) | 15 (13.8%) | 80 (21.2%) |  |
| b. Dead | 4 (2.0%) | 3 (3.8%) | 1 (0.9%) | 8 (2.1%) |  |
| - Home quarantine | 0 | 1 (33.3%) | 0 | 1 (12.5%) |  |
| - Institutional quarantine/<br>Hospitalization | 4 (100%) | 2 (66.7%) | 1 (100%) | 7 (87.5%) |  |
| **8. Vaccination Status** |  |  |  |  | 0.288 |
| Not vaccinated | 18 (9.2%) | 05 (6.2%) | 13 (11.8%) | 36 (9.3%) |  |
| Vaccinated | 178 (90.8%) | 75 (93.8%) | 97 (88.2%) | 350 (90.7%) |  |
| a. Vaccinated with one dose | 13 (7.3%) | 2 (2.7%) | 2 (2.1%) | 17 (4.9%) |  |
| b. Vaccinated with two doses | 144 (80.9%) | 52 (69.3%) | 69 (71.1%) | 265 (75.7%) |  |
| c. Vaccinated with precautionary dose (Booster dose) | 21 (11.8%) | 21 (28.0%) | 26 (26.8%) | 68 (19.4%) |  |

While 25.7% of XBB.1.16* cases were institutionally quarantined or hospitalized, most cases (15/71, 21.1%) were admitted for reasons other than COVID-19. The mean duration of hospital stay for XBB.1.16* cases was 6.6 ± 3.9 days. Among the home isolated XBB.1.16* cases, 96.1% required conservative treatment. On the other hand, 64.8% of hospitalized cases were given conservative treatment and 33.8% of cases required supplemental oxygen therapy. Out of all the XBB.1.16* cases, 97.5% recovered from the disease, while 2.5% succumbed to the disease. However, there was no significant difference in the survival of XBB.1.16 and other Omicron cases.

Covishield (ChAdOx1nCoV-19 Corona Virus Vaccine) (81.9%) was the most common vaccine administered, followed by Covaxin (BBV152A-a whole inactivated virus-based COVID-19 vaccine) (5.7%). Among the 386 cases, 350 (90.7%) were vaccinated with at least one dose of the COVID-19 vaccine, and 36 (9.3%) were unvaccinated. Most unvaccinated individuals were in the age group of 0 to 10 years (44.4%) and were not offered vaccination as a part of the vaccination policy in the country.

It is important to note that the study did not find any significant difference in the clinical presentations of XBB.1.16 and XBB.1.16.1 cases. Further, the clinical symptoms of XBB.1.16* cases resembled those of other co-circulating Omicron lineage infected cases and caused mild disease.

### 3.4. Characteristics of death cases during the study

A total of eight cases (2.1%) died during the study. Six out of eight (75%) deaths occurred in the age group of 60 years and above (75%) (***Table 5, Figure 8***). Presence of comorbidity was seen in five (62.5%) cases (***Figure 9***). Vaccination with at least one dose of vaccine was present in 87.5% of cases. However, there was no significant difference in vaccination status of survived and death cases. Seven out of eight cases (87.5%) were hospitalized of which six cases (85.7%) needed oxygen therapy. Out of the total number of cases, five cases belonged to the elderly age group (60 years and above) and had underlying comorbidities. In one case, the individual belonged to the age group of 0 to 9 years and the cause of death was drowning, while COVID-19 was an incidental finding. For the remaining two cases, the cause of death could not be determined.

**Table 5:** Characteristics of survived and dead cases in the study

| Characteristics | Survived | Dead | *p*-value |
| --- | --- | --- | --- |
| **1. Gender-wise distribution** |  |  | 1.000 |
| Male | 198 (52.4%) | 4 (50%) |  |
| Female | 180 (47.6%) | 4 (50%) |  |
| **2. Age-wise distribution** |  |  | < 0.001 |
| 0 to 9 | 17 (4.5%) | 1 (12.5%) |  |
| 10 to 19 | 33 (8.7%) | 0 |  |
| 20 to 29 | 88 (23.3%) | 0 |  |
| 30 to 39 | 86 (22.8%) | 1 (12.5%) |  |
| 40 to 49 | 48 (12.7%) | 0 |  |
| 50 to 59 | 50 (13.2%) | 0 |  |
| 60 and above | 56 (14.8%) | 6 (75.0%) |  |
| **3. Vaccination status** |  |  | 0.755 |
| Vaccinated | 343 (90.7%) | 7 (87.5%) |  |
| Not vaccinated | 35 (9.3%) | 1 (12.5%) |  |
| **4. Symptom status at the time of acute infection** |  |  | 0.431 |
| Asymptomatic | 25 (6.6%) | 1 (12.5%) |  |
| Symptomatic | 353 (93.4%) | 7 (87.5%) |  |
| **5. Underlying comorbid conditions** |  |  | < 0.001 |
| Absent | 321 (84.9%) | 3 (37.5%) |  |
| Present | 57 (15.1%) | 5 (62.5%) |  |
| **6. Type of quarantine** |  |  | < 0.001 |
| Home isolated | 298 (78.8%) | 1 (12.5%) |  |
| Hospitalized | 80 (21.2%) | 7 (87.5%) |  |
| **7. Oxygen requirement among hospitalized cases** |  |  | 0.004 |
| Oxygen therapy not needed | 55 (68.7%) | 1 (14.3%) |  |
| Oxygen therapy needed | 25 (31.3%) | 6 (85.7%) |  |

**Figure 8:**
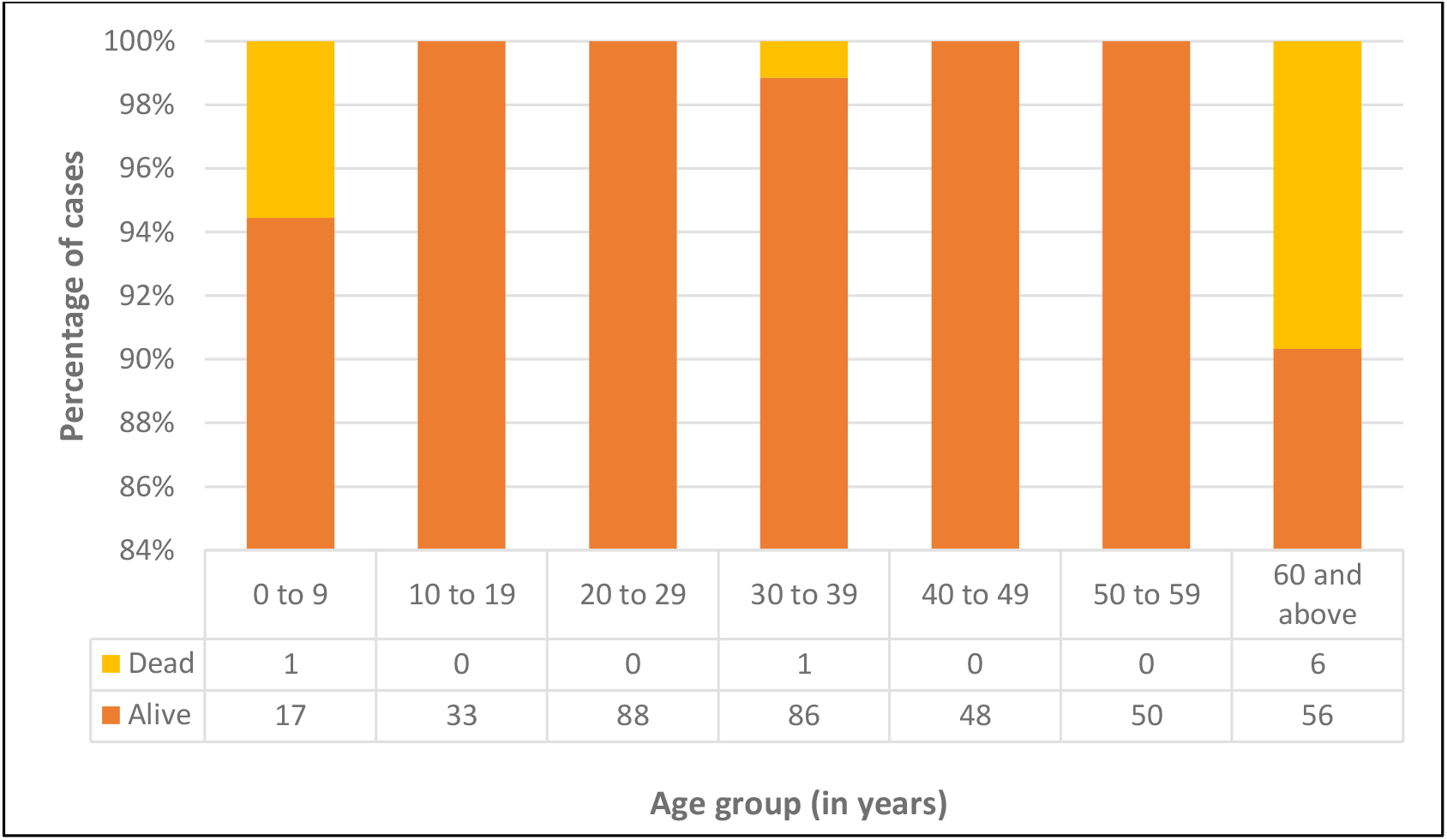
Age-wise distribution of survived and dead cases

**Figure 8:**
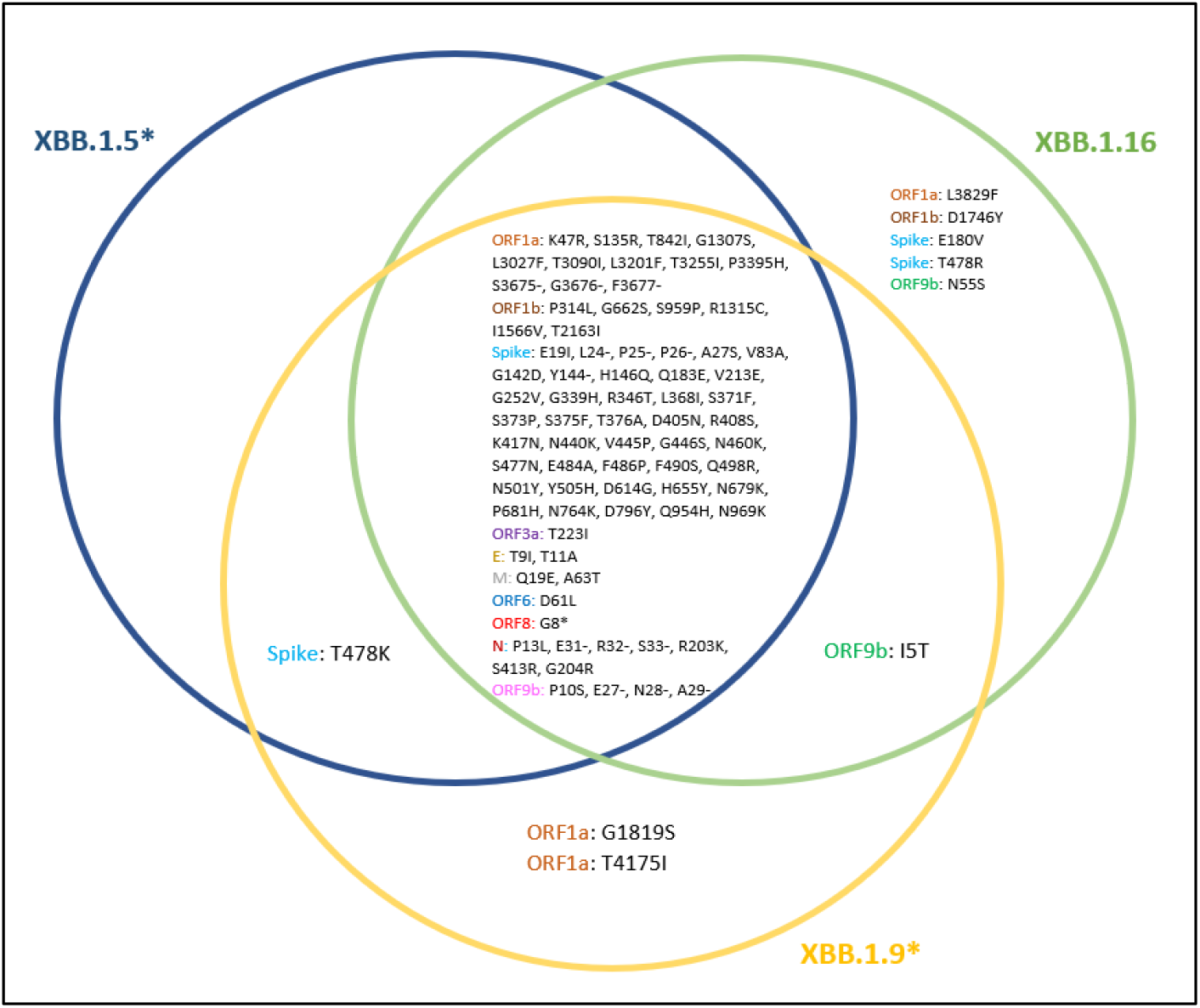
Mutations unique to XBB.1.16 lineage and those shared with XBB.1.5* and XBB.1.9* lineages

**Figure 9:**
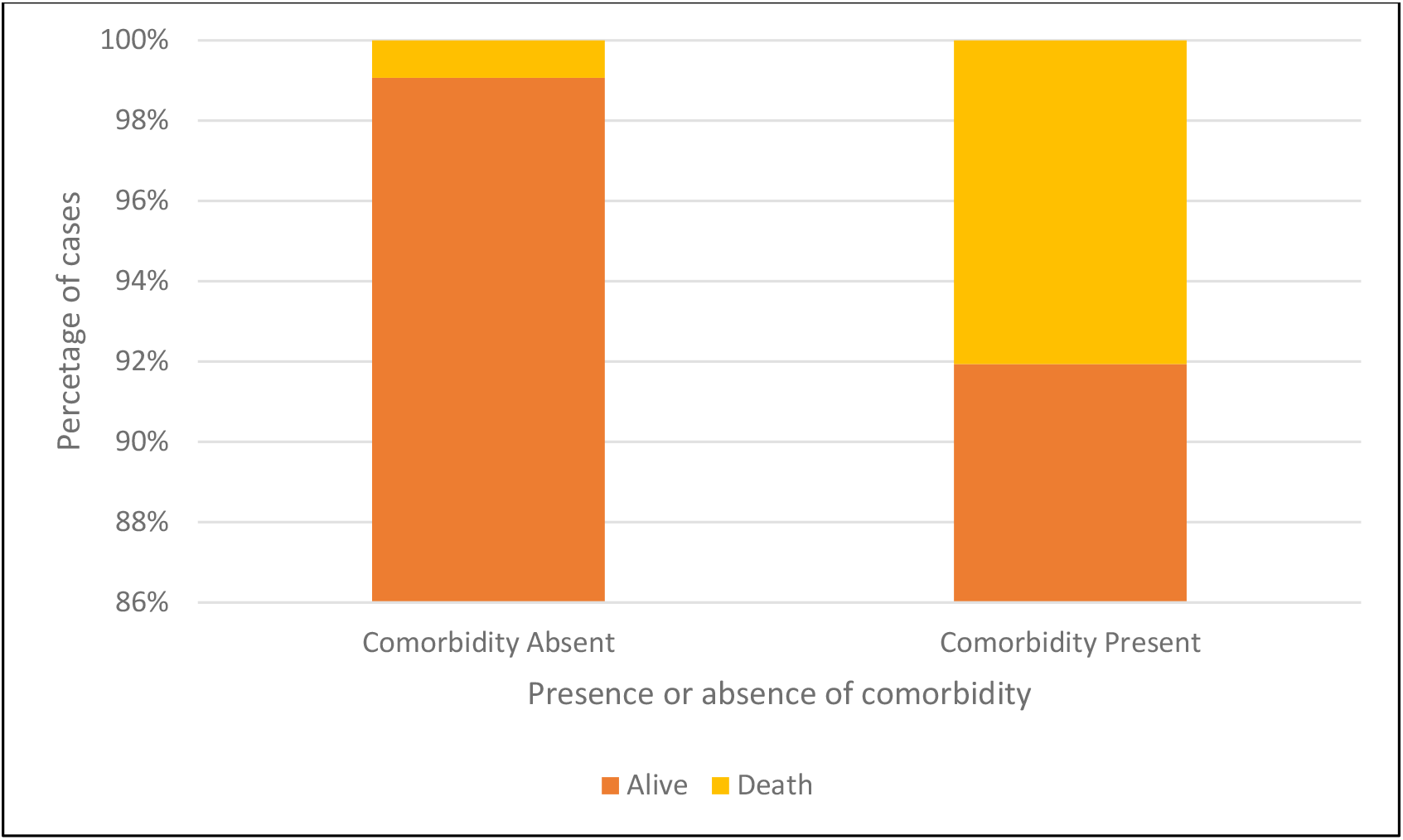
Presence or absence of comorbidity among survived and dead cases

## 5. DISCUSSION

Despite an overall decrease in newly reported COVID-19 cases worldwide, the WHO South-East Asia Region and the Eastern Mediterranean Region are experiencing a significant increase in the number of COVID-19 cases and deaths over the past 28 days (from 13^th^ March 2023 to 9^th^ April 2023). The countries in WHO South-East Asia region reporting highest number of cases include India, Indonesia, and Thailand. While the Variant of Interest (VOI), XBB.1.5 SARS-CoV-2 variant, accounts for most cases (47.9%) globally (as on 9^th^ April 2023) [16]. Since its identification on 4^th^ March 2023, the XBB.1.16* lineage has already spread to 31 countries, with India reporting the highest percentage of sequences (32.47%), followed by Brunei (4.50%) and Singapore (3.21%) [17]. Worldwide, the cumulative prevalence of XBB.1.16 is less than 0.5%, while its cumulative prevalence in India is 2% (on 17^th^ April 2023) [18]. With the current relative growth advantage of 62% (95% Confidence Interval – 56 to 68%) in India, the lineage has already been found in sequences from 15 Indian states and Union Territories (till 10^th^ April 2023) [19] and accounts for 49% (95% Confidence Interval – 46 to 51%) of SARS-CoV-2 sequences over the past 60 days [18]. On 22^nd^ March 2023, the World Health Organization (WHO) added XBB.1.16 to the Variant Under Monitoring (VUM) list along with BQ.1, BA.2.75, CH.1.1, XBB and XBF [20]. Further, on 11^th^ April 2023, Nextstrain/Nextclade (https://github.com/nextstrain/ncov) elevated XBB.1.16 as new clade 23B [21]. On 17^th^ April 2023, XBB.1.16 was added to the WHO list of VOIs [22].

XBB.1.16 is an XBB.1 sub-lineage characterized by unique mutations in the genome’s spike and open reading frame regions. It also shares mutations with XBB.1.5 and XBB.1.9 lineage (***Figure 8***). Specifically, the spike mutations include S: E180V in the N-terminal domain and S: T478R, S: S486P in the receptor binding domain (RBD) of the spike protein, and the mutations in the open reading frames include ORF1a: L3829F, ORF1b: D1746Y, ORF9b: I5T and ORF9b: N55S [9]. According to a recent study describing the virological features of XBB.1.16, the binding affinity of XBB.1.16 RBD to human angiotensin-converting enzyme 2 (ACE2) receptor is higher than XBB.1 but lower than XBB.1.5. The spike mutations have varying effects on infectivity, with S: T478R substitution significantly increasing the infectivity and S: E180V decreasing it. Furthermore, it is also showed that XBB.1.16 is antigenically different from XBB.1.5, thereby contributing to its increased fitness over XBB.1.5. Similar combination mutations have also been observed in BA.5, BA.2.75 and XBB.1, suggesting XBB.1.16 possibly follows a similar evolutionary pattern [23].

Though a high percentage of the study population received at least one dose of vaccine (90.7%), breakthrough infections were observed frequently in the study. In-vitro neutralization assays demonstrated that XBB.1.16 exhibited strong resistance to breakthrough infections by sera from both BA.2 (18-fold) and BA.5 (37-fold) lineages. Furthermore, XBB.1.16 was as sensitive to the convalescent sera of XBB.1-infected hamsters as XBB.1 and XBB.1.5. Like other XBB sublineages, XBB.1.16 was found to be resistant to six clinically available monoclonal antibodies, with only Sotrovimab retaining antiviral activity against XBB sublineages. Consequently, due to its robust resistance to various anti-SARS-CoV-2 antibodies, XBB.1.16 is more immune evasive when compared to XBB.1 and XBB.1.5 [23].

In addition to the spike proteins, the non-spike proteins of the virus have significant roles in regulating the immune response, controlling viral transcription and in viral pathogenesis [24]. For example, the XBB variant has retained the mutation T9I (99.70% in BA.1) in its envelope (E) protein, found earlier in Omicron sub-variants. It has also gained a new negative mutation T11A at a high frequency (90.52%). These changes in the envelope protein are believed to reduce the virulence and pathogenicity of the variant [25, 26]. Therefore, it is possible that individuals infected with XBB.1.16 lineage, like those infected with other Omicron lineages, experienced a mild disease, despite exhibiting symptoms. Similarly, accessory proteins like ORF9b play a significant role in viral pathogenesis by reducing the host antiviral response [24]. The ORF9b gene of the XBB.1.16 lineage has two distinct substitutions, I5T (Isoleucine → Threonine) and N55S (Asparagine → Serine) [9]. ORF9b suppresses and antagonizes the type I interferon response (IFN-I) by interacting with TOM70 and by targeting multiple signalling pathways like the RIG-I-MAVS-dependent IFN signalling pathway, leading to innate immune suppression [24,25,27]. The mutation N55S lies in the ORF9b region (43-78 residues) that interacts with TOM70, while the I5T mutation is located in the N terminal site, which helps to stabilize the TOM70-ORF9b structure. However, the effect of these mutations remains uncertain [24]. Therefore, it is essential to conduct further studies to determine the effects of these unique mutations, as they could potentially impact the virus’s interaction with the host immune system and contribute to disease pathogenesis.

## 6. CONCLUSION

The study reveals that XBB.1.16* lineage has become the most predominant SARS-CoV-2 lineage in India. The study also shows that the clinical features and outcome of XBB.1.16* cases were similar to those of other co-circulating Omicron lineage infected cases in Maharashtra, India. The XBB.1.16 variant did not cause severe infections just like other Omicron sub-lineages. However, its increased transmissibility and immune evasive properties are alarming. Moreover, due to the increased growth efficiency, XBB.1.16 variant is progressively replacing all other co-circulating lineages in India. This study underlines the importance of a prompt assessement of clinical characteristics and outcomes following the rapid identification of a new SARS-CoV-2 lineage (XBB.1.16 variant in this study) as well as the immediate dissemination of genomic data to public databases, such as GISAID. The first action mentioned above is important for clinical genomic surveillance and the second is essential in detection and naming of new lineages. The results of both measures mentioned previously, provides a prompt actionable evidence to aid policymakers in making informed public health decisions and interventions (<u>14042023XBB.1</u> (who.int)) [28].

## Supporting information

https://epicov.org/epi3/epi_set/230419bg?main=true

## Data Availability

All data produced in the present work are contained in the manuscript

https://epicov.org/epi3/epi_set/230419bg?main=true

## ACKNOWLEDGEMENT

We gratefully acknowledge all data contributors, i.e., the Authors and their Originating laboratories responsible for obtaining the specimens, and their Submitting laboratories for generating the genetic sequence and metadata and sharing via the GISAID Initiative, on which this research is based.

The authors wish to thank Dr Akshay Karyakarte, Assistant Professor, Department of Microbiology, Seth GS Medical College and KEM Hospital, Mumbai, for providing intellectual assistance and technical help in writing and editing the manuscript.

We also thank Mrs Poonam Pacharne, Mr Vishal Rajput, and Miss Riyakshi Rajkhowa from Byramjee Jeejeebhoy Government Medical College and Sassoon General Hospitals, Pune, for their technical help during the study.

